# Planner-Executor Style Multimodal Agentic System to Answer Patient Questions in Lung Cancer Screening CT

**DOI:** 10.64898/2026.10.02.26364620

**Authors:** Koharu Sakiyama, Adrian Serapio, Meng Ye, Ali Nowroozi, Masha Bondarenko, Yu-Fu Wu, Kang Qi, Jonathan Liu, Maya Vella, Yannan Yu, Tician Schnitzler, Alison S Rustagi, Jae Ho Sohn

## Abstract

**Background:** Accurate patient understanding of lung cancer screening (LCS) results is critical for engagement and follow-up adherence, given the currently low screening uptake. While artificial intelligence (AI) systems show promise in facilitating patient communication, its effective use in clinical settings requires appropriate invocation of imaging tools and self-regulation by deferring certain questions to physicians. To this end, we developed and evaluated a planner-executor style multimodal agentic system to answer simulated patient questions about LCS CT results.

**Methods:** This retrospective study utilized 116 LCS CT reports and images collected from a tertiary academic hospital. The system employed a multi-agent architecture (planner and executor) to mimic clinical reasoning. We developed a large language model (LLM)-based question generation framework to prepare a comprehensive set of 699 simulated patient questions, categorized as answerable and defer-to-doctor to assess self-regulation. Performance was evaluated on tool-calling, self-regulation accuracy (ability to correctly defer out-of-scope questions to a human provider) assessed by LLM-as-a-judge, and clinical quality assessed by a reader performance study with four physicians evaluating a subset of 100 responses.

**Results:** The system demonstrated high overall tool-calling accuracy of 93.1% (646/694; 95% CI: 91.2%, 95.0%) and self-regulation accuracy of 92.6% (462/499; 90.2%, 94.8%). For answerable questions, the percentage of responses receiving perfect 5.0 scores across all readers included 78.9% (180/228) for clinical accuracy (inter-rater agreement: Gwet’s AC2=0.92), and 73.2% (167/228) for patient understandability (0.86). For defer-to-doctor questions, the percentage of responses included 77.3% (133/172) for clinical accuracy (0.80) and 70.3% (121/172) for patient understandability (0.78).

**Conclusions:** The planner-executor style multi-modal agentic system reliably answers patient-specific questions about LCS. Its high tool-calling and self-regulation performance demonstrates its potential as a safe and effective digital communication facilitator in LCS programs.

## INTRODUCTION

Effective patient communication in lung cancer screening (LCS) is critical for improving the historically low rates of screening uptake and follow-up adherence [1], [2]. Patient engagement with medical information has surged following the 21st Century Cures Act, which granted patients near-immediate access to electronic health records, including radiology reports [3]. While this transparency empowers patients, they frequently review complex findings before meeting with referring providers [4]. In LCS specifically, reports often contain technical classifications, such as Lung-RADS categories and incidental nodule measurements, that are crucial for management but frequently misinterpreted by laypersons as worrisome diagnoses.

In the absence of immediate clinical guidance, this "information gap" often leads to confusion regarding medical terminology, heightened anxiety, and an increased volume of inquiries to clinical staff. Enhancing patient understanding of LCS results is essential to maintaining engagement and ensuring timely intervention, which ultimately improves outcomes. However, the clinical capacity to address these questions is increasingly constrained by time and workforce demands. This gap highlights an urgent need for scalable, automated strategies that can support patient understanding and facilitate effective communication while minimizing the administrative burden on clinicians.

Advancements in generative artificial intelligence (AI), particularly large language models (LLMs), have demonstrated significant potential in simplifying radiology reports and facilitating patient understanding [5], [6]. However, radiological communication requires multimodal, domain-specific interpretation of specialized data that often exceeds the capabilities of general-purpose models. While numerous computer vision and natural language processing tools have been developed for radiology-specific tasks [7], [8], offering opportunities to enhance automated systems, effective integration of these tools requires models to accurately identify and invoke appropriate tools. Furthermore, some clinical inquiries such as prognostic or treatment-related questions often need to be deferred to clinical expertise to ensure reliability and accuracy in the medical setting.

To this end, a "planner-executor" framework can be employed to mimic clinical reasoning. This architecture first decomposes a patient’s query into logical sub-tasks before retrieving relevant report data or clinical guidelines. This separation of logic and execution allows for a dedicated verification step, ensuring that the final response is both linguistically coherent and clinically grounded. To address the need for reliable automated communication, we developed and evaluated a planner-executor agentic AI system with integrated tool-calling and self-regulatory capabilities, systematically validated across a diverse cohort of LCS CT reports.

## METHODS

### Data Acquisition

CT reports and scans were retrospectively collected under approval of the Institutional Review Board, with informed consent waived. A total of 1,052 LCS CT reports and scans between 18 October 2017 and 19 July 2023 were obtained from a tertiary academic center. Using pattern matching, reports that did not contain ‘Lung-RADS’, assigned classifications of 0, or with incomplete imaging were excluded. From the remaining dataset, 116 cases were selected by stratified sampling, with sample size determined by the power analysis detailed in the Appendix. Stratification was performed to match the Lung-RADS category distribution observed at our institution, provided in the Appendix. Six additional reports were sampled to create a prompt development set. Patient characteristics are detailed in Table 1.

**Table 1:**
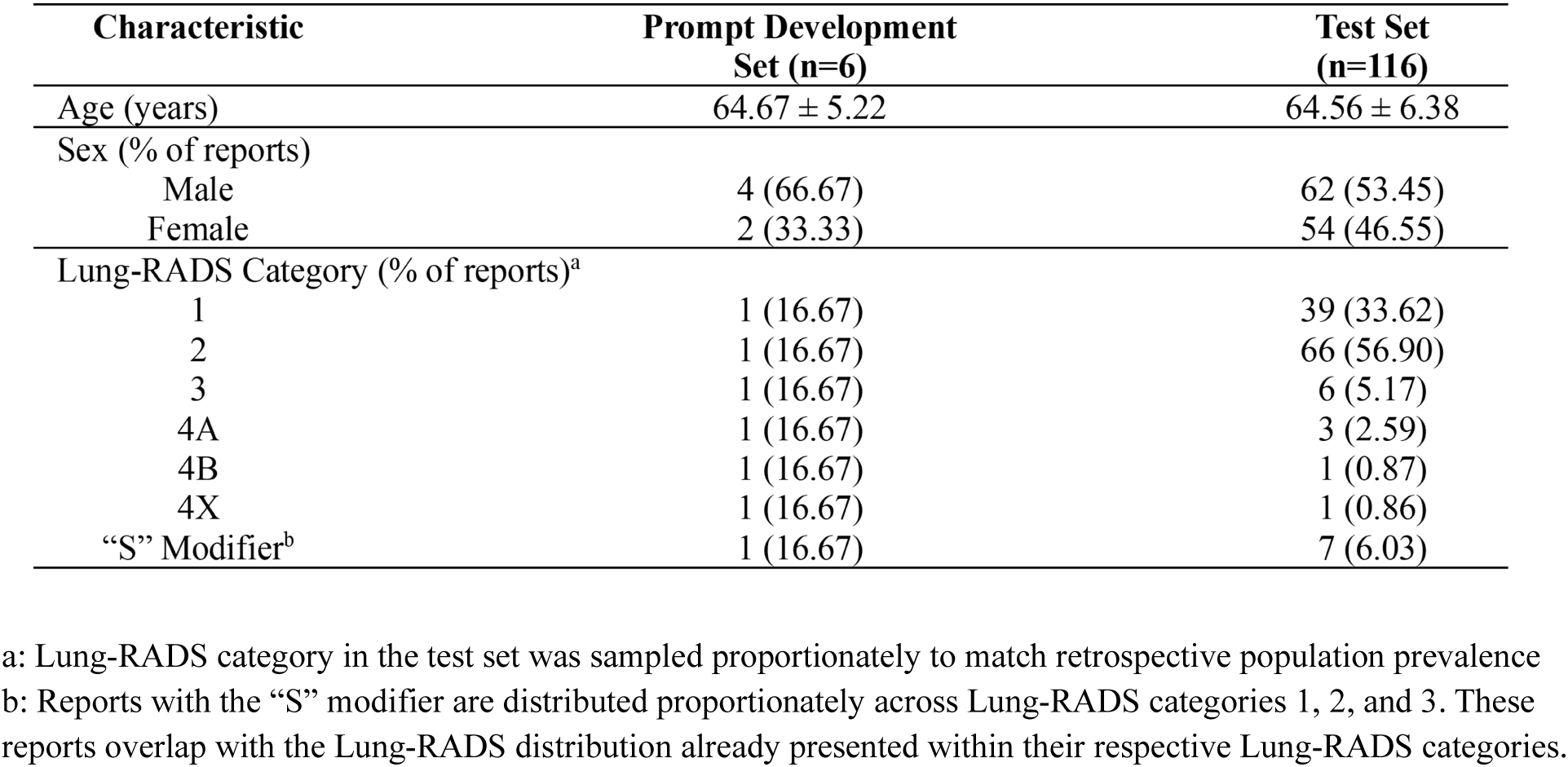
Patient demographics and characteristics of lung cancer screening CT reports used for generating simulated patient questions and agentic system evaluation.

| Characteristic | Prompt Development Set (n=6) | Test Set (n=116) |
| --- | --- | --- |
| Age (years) | 64.67 ± 5.22 | 64.56 ± 6.38 |
| Sex (% of reports) |  |  |
| Male | 4 (66.67) | 62 (53.45) |
| Female | 2 (33.33) | 54 (46.55) |
| Lung-RADS Category (% of reports)<sup>a</sup> |  |  |
| 1 | 1 (16.67) | 39 (33.62) |
| 2 | 1 (16.67) | 66 (56.90) |
| 3 | 1 (16.67) | 6 (5.17) |
| 4A | 1 (16.67) | 3 (2.59) |
| 4B | 1 (16.67) | 1 (0.87) |
| 4X | 1 (16.67) | 1 (0.86) |
| “S” Modifier<sup>b</sup> | 1 (16.67) | 7 (6.03) |

a: Lung-RADS category in the test set was sampled proportionately to match retrospective population prevalence
b: Reports with the “S” modifier are distributed proportionately across Lung-RADS categories 1, 2, and 3. These reports overlap with the Lung-RADS distribution already presented within their respective Lung-RADS categories.

### Agentic System Architecture

Our agentic system is composed of two agents, a planner and executor. The planner receives as input the question and extracted CT report (comparison, findings and impression sections), and outputs a structured step-by-step plan, specifying tool invocation and determining whether the query is answerable or should be deferred to a physician. The executor implements the plan, invoking imaging tools, referring to Lung-RADS v2022 guidelines [9], and reviewing the CT report to synthesize a patient-friendly response (Fig. 1). Both agents are powered by a HIPAA-compliant GPT-4o [10], which has demonstrated success in related medical agentic systems [11], [12].

**Figure 1.**
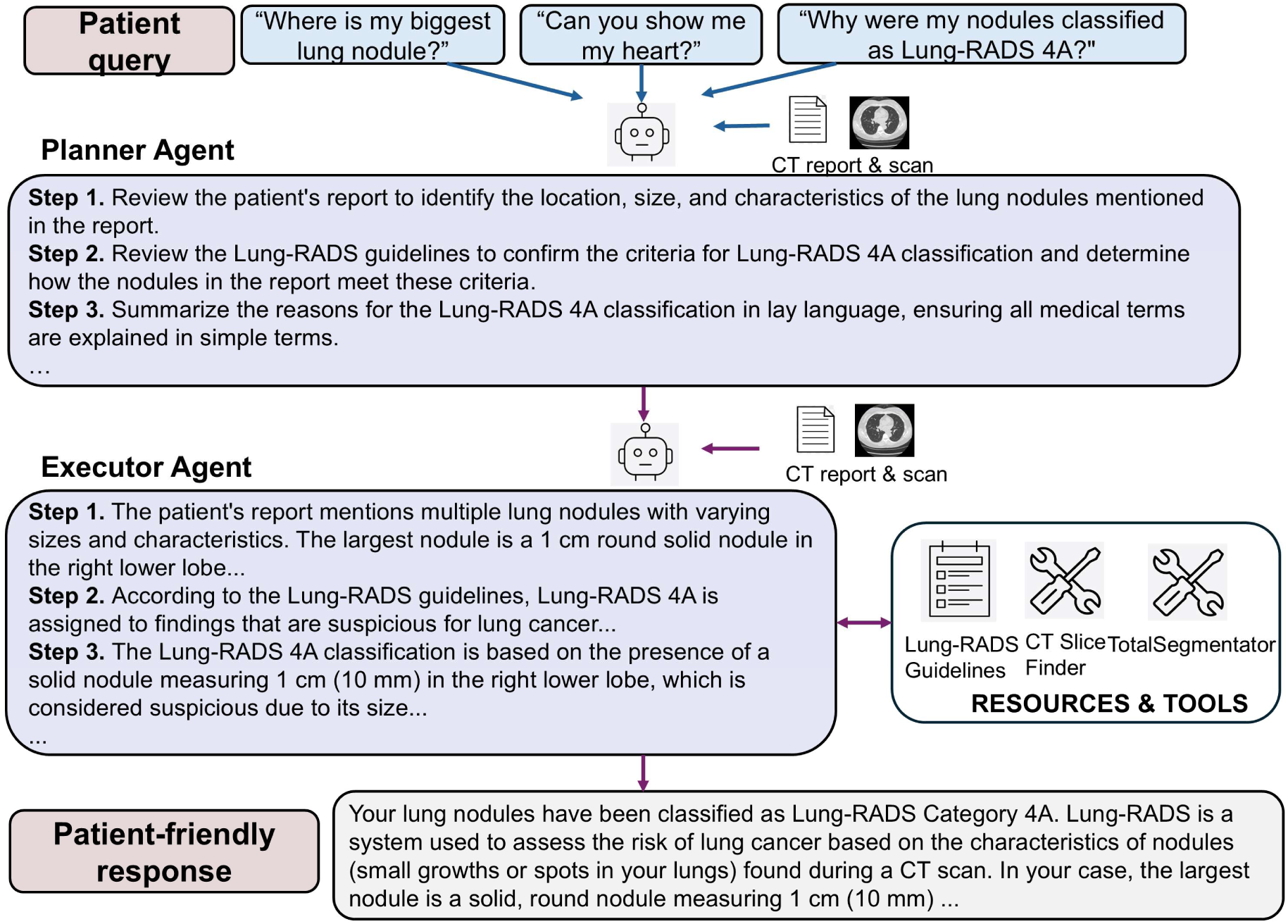
Agentic system architecture. The multimodal agentic system employs a multi-agent architecture consisting of a planner and an executor agent, both of which have access to the patient’s CT scan and report. The planner agent interprets the patient query and outputs a structured step-by-step plan, and the executor agent monitors and implements this plan, invoking imaging tools, referring to Lung-RADS v2022 guidelines, and reviewing the CT report to synthesize a patient-friendly response.

To expand the system’s capability to process 3D imaging data, two specialized tools were integrated: a Python-based parsing function to retrieve CT images referenced in the report, and TotalSegmentator [13], an open-source segmentation model for visualization of relevant anatomical structures. Preprocessing steps were applied prior to running TotalSegmentator, including the selection of the largest CT series and the conversion of DICOM files to NIfTI format to ensure compatibility. The organ segmentation mask with the largest area was then overlaid onto the original NIfTI image to allow visualization of the target anatomy segmentation.

Predefined instructions in the agent prompts emphasized self-regulation by limiting the system to invoking tools, referring to the Lung-RADS guidelines and CT report, and simplifying medical terms. Various combinations of zero-shot [14] and few-shot [15] prompting strategies were applied to both agents, and a single-agent system was developed as a baseline. Prompts for both agents (Supplementary Fig. 1) were iteratively refined using a set of 36 questions derived from reports in the prompt development set through evaluating tool-calling accuracy, self-regulation accuracy with human-labeled ground truths, and feedback from two medical experts.

### LLM-assisted Question Generation

To evaluate our agentic system, we generated 699 simulated patient questions using a HIPAA-compliant GPT-4o, based on the sampled CT reports and informed by prior work on thematic analysis of patient questions in LCS (Fig. 2) [16]. The prepared questions were then categorized into "answerable" and "defer-to-doctor" categories. The system was instructed to defer if questions required patient-specific history, prognosis, or lacked sufficient information.

**Figure 2.**
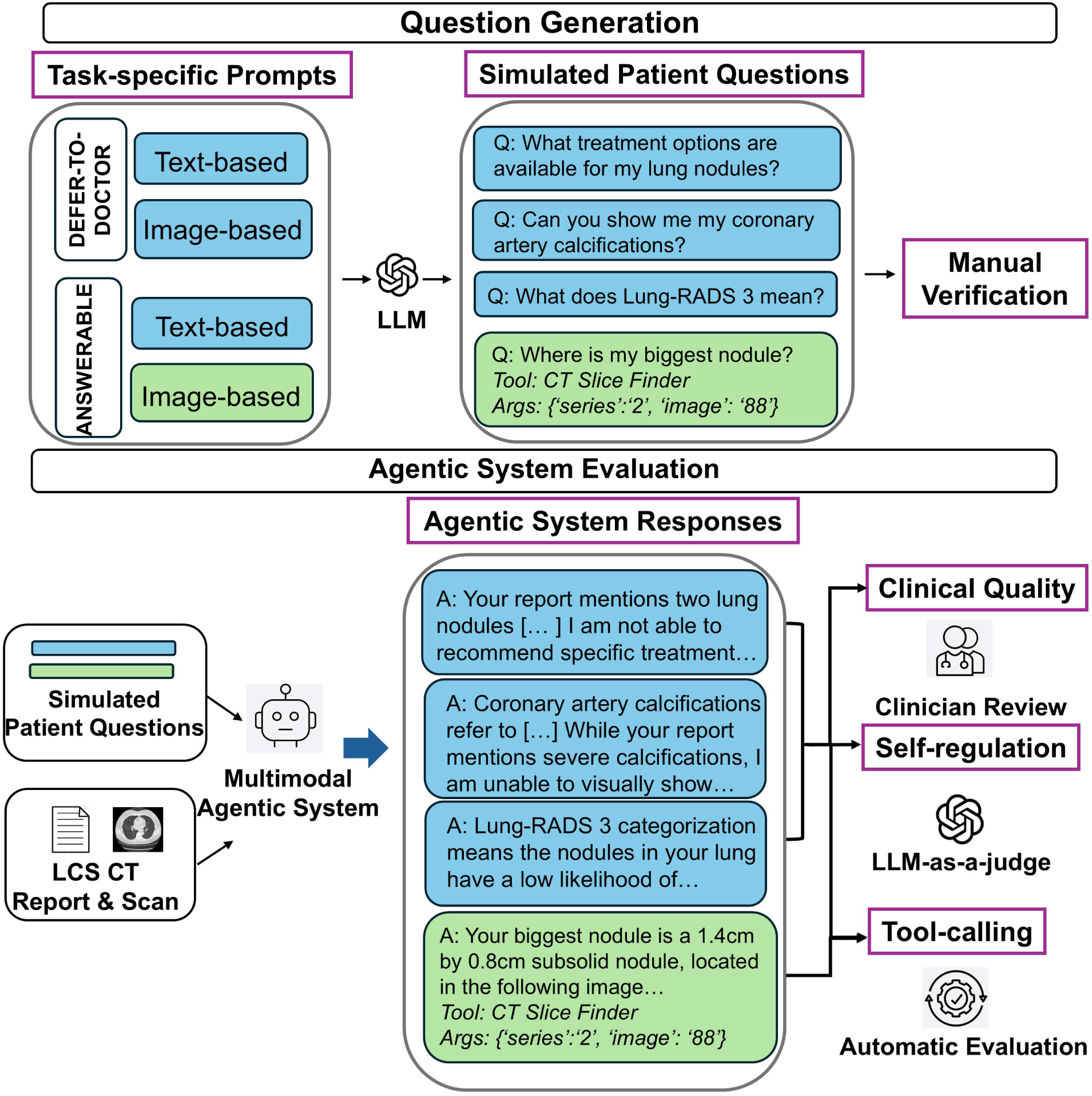
Workflow for large language model (LLM)-based question generation and multimodal agentic system evaluation. Task-specific prompts were used to generate a comprehensive set of simulated patient questions with LLM assistance and manually verified. Questions were broadly categorized as answerable or defer-to-doctor. Agentic system responses to these questions were evaluated using three metrics: tool-calling accuracy assessed through automated evaluation, self-regulation accuracy assessed by an LLM-as-a-judge, and clinical quality evaluated in a reader performance study by four physicians on a subset of 100 responses.

Answerable questions were sub-categorized into image-based questions requiring tool-calling and text-based questions addressed using the CT report or Lung-RADS guidelines. Image-based questions were further divided into tasks of localizing a finding or anatomical structure mentioned in the report, corresponding to each tool. Text-based questions covered tasks including Lung-RADS category interpretation, comparisons using information in the CT report, and providing lay language explanation of medical terms for answerable questions, and patient-specific treatment, follow-up of incidental findings, cause of finding, lifestyle changes, and cancer staging for defer-to-doctor questions. Question generation prompts (Supplementary Fig. 2) were iteratively refined using the six reports in the prompt development set through clinician feedback. All questions, tool calls, and tool parameters generated in the process were additionally verified to ensure accuracy and quality. Table 2 provides examples and quantification of each question type.

**Table 2:** Question types and descriptions of the simulated patient question set.

| Type | Category | Description | Examples | Tool Call | Quantity in Prompt Development Set | Quantity in Test Set |
| --- | --- | --- | --- | --- | --- | --- |
| Answerable | Image-based | Questions requiring tool call to retrieve relevant CT slice or segment anatomical structure | - *Can you show me my pulmonary nodule?*<br>- *Where is the right lower lobe of my lungs, where my nodule was mentioned?* | Y | 10 | 195 |
|  | Text-based | Questions requiring interpretation of Lung-RADS guidelines, reasoning about comparisons based on CT report, or lay language explanation of medical terms | - *What does Lung-RADS 4A mean?*<br>- *Can you explain what mild centrilobular emphysema is?*<br>- *What differences are there compared to my previous scan?* | N | 14 | 276 |
| Defer-to-doctor | Image-based | Questions requiring visualization of CT slice, annotations, or anatomical structures beyond the capability of provided tools | - *Can you show me my coronary artery calcifications?*<br>- *Can you show me where the atelectasis is on my scan?* | N | 6 | 112 |
|  | Text-based | Questions requiring patient-specific medical history such as cause of disease, prognosis, lifestyle changes, patient-specific treatment | - *What could have caused me to develop nodules in my lungs?*<br>- *What can I do to make sure my lung nodules don't grow?*<br>- *What should I do to manage the mild emphysema mentioned in the report?* | N | 6 | 116 |

### Automated Evaluation for Tool-calling

Tool-calling accuracy was assessed using accuracy and F1 scores. True positives were defined as cases where the system correctly invoked the reference standard tool with correct parameters. Per-tool accuracy was calculated for each imaging tool, with overall accuracy computed as the proportion correct across all tool classes. Of the 699 responses, five were excluded from tool-calling analysis due to ambiguity in the reference standard. Clinician input, blind to model output, determined that both tool invocation and non-invocation could be considered clinically appropriate based on the question phrasing. These cases were excluded to avoid introducing bias into accuracy calculations.

### LLM-as-a-judge for Self-Regulation

We implemented an LLM-as-a-judge to evaluate the self-regulation accuracy of the agentic system in a scalable manner. Building on existing work utilizing LLMs as evaluators in the medical domain [17], [18], we assessed the performance of LLM-based evaluation by comparing its alignment to human evaluation by the author on a subset of 105 responses across various LLMs at a temperature of 0.0 (Supplementary Fig. 3). The LLM demonstrating the highest alignment, GPT-4o (alignment 0.99 accuracy), was selected for the full evaluation. The LLM classified the 499 text-based responses as “correct” or “incorrect” based on alignment with their intended categories (answerable or defer-to-doctor). Self-regulation accuracy was assessed using precision, recall, and F1 scores. Overall deferral accuracy was calculated as the proportion of correct labels across both categories. Prompts are provided in Supplementary Fig. 4.

### Reader Performance Study

We then conducted a reader performance study with a post-graduate year 5 radiologist in-training, two board-certified radiologists (5 and 6 years of post-cardiothoracic fellowship experience respectively), and a thoracic surgeon (8 years post-graduate experience). A random subset of 100 text-based responses, proportionately sampled across answerable and defer-to-doctor types, was evaluated for clinical quality. Responses were rated on a 1-5 Likert scale for factuality, conciseness, and patient understandability. Additionally, answerable questions were rated for comprehensiveness and defer-to-doctor questions for relevance to the query. Sampling details and evaluation criteria are provided in Supplementary Fig. 5-6.

### Statistical Analysis

Nonparametric bootstrapping based on repeated random sampling was used to calculate 95% confidence intervals. McNemar’s test [19] with Bonferroni correction for multiple comparisons was used to compare accuracy of multi-agent variations against the single-agent baseline, with adjusted significance level P ≤ 0.0125 (α=.05/4). Gwet’s agreement coefficient weighted for ordinal ratings (AC2) [20] was used to measure inter-rater agreement in the reader study. Similarity in the proportion of perfect 5.0 scores across question types was evaluated using Fisher’s exact test (P < 0.05) [21].

## RESULTS

### Automated Evaluation for Tool-calling

The agentic system’s ability to accurately select the appropriate tool among the CT slice retrieval tool, segmentation tool, or no tool use was evaluated for all questions, with results summarized in Table 3. Compared to the single-agent baseline, which achieved overall tool-calling accuracy of 78.5% (545 of 694; 95% CI: 75.5%–81.4%), all variations of the planner-executor multi-agent architecture demonstrated significantly higher accuracy (all P < .001).

**Table 3:** Tool-calling accuracy with varying prompting strategies in the planner and executor agents.

| Planner Prompt | Executor Prompt | Tool Name | Accuracy (%) (95% CI) | F1 Score (95% CI) | Overall Accuracy (%) (95% CI) | Adjusted P-value<sup>a</sup> |
| --- | --- | --- | --- | --- | --- | --- |
| Few-shot | Few-shot | CT Slice Finder | 97.7 (678/694)<br>(96.5–98.7) | 0.906<br>(0.855–0.947) | 93.1 (646/694)<br>(91.2–95.0) | <0.001 |
|  |  | TotalSegmentator | 95.4 (662/694)<br>(93.8–96.8) | 0.878<br>(0.834–0.917) |  |  |
|  |  | No Tool Call | 93.4 (648/694)<br>(91.5–95.1) | 0.952<br>(0.937–0.965) |  |  |
| Zero-shot | Few-shot | CT Slice Finder | 96.7 (671/694)<br>(95.2–98.0) | 0.864<br>(0.804–0.916) | 89.0 (618/694)<br>(86.6–91.4) | <0.001 |
|  |  | TotalSegmentator | 91.5 (635/694)<br>(89.5–93.5) | 0.776<br>(0.717–0.828) |  |  |
|  |  | No Tool Call | 90.5 (628/694)<br>(88.3–92.7) | 0.931<br>(0.913–0.947) |  |  |
| Few-shot | Zero-shot | CT Slice Finder | 96.5 (670/694)<br>(95.1–97.8) | 0.864<br>(0.804–0.914) | 92.1 (639/694)<br>(90.1–94.1) | <0.001 |
|  |  | TotalSegmentator | 95.5 (663/694)<br>(93.9–97.0) | 0.882<br>(0.838–0.921) |  |  |
|  |  | No Tool Call | 92.5 (642/694)<br>(90.5–94.4) | 0.945<br>(0.930–0.959) |  |  |
| Zero-shot | Zero-shot | CT Slice Finder | 96.5 (670/694)<br>(95.1–97.8) | 0.862<br>(0.802–0.912) | 88.8 (616/694)<br>(86.3–91.1) | <0.001 |
|  |  | TotalSegmentator | 91.5 (635/694)<br>(89.3–93.5) | 0.770<br>(0.709–0.824) |  |  |
|  |  | No Tool Call | 89.6 (622/694)<br>(87.3–91.8) | 0.925<br>(0.906–0.941) |  |  |
| Single-agent baseline |  | CT Slice Finder | 90.8 (630/694)<br>(88.5–92.8) | 0.695<br>(0.618–0.762) | 78.5 (545/694)<br>(75.5–81.4) | N/A |
|  |  | TotalSegmentator | 84.9 (589/694)<br>(82.1–87.5) | 0.688<br>(0.630–0.742) |  |  |
|  |  | No Tool Call | 79.4 (551/694)<br>(76.4–82.3) | 0.833<br>(0.804–0.858) |  |  |

a: McNemar’s test with Bonferroni correction for multiple comparisons was used to compare accuracy of multi-agent variations against the single-agent baseline, with adjusted significance level $P \leq 0.0125$ ( $\alpha=.05/4$ ).

Using the multi-agent architecture, we additionally compared combinations of zero-shot and few-shot prompting for both the planner and executor. The highest-performing planner-executor system, using few-shot prompting for both agents, achieved an overall tool-calling accuracy of 93.1% (646 of 694; 95% CI: 91.2–95.0%). Performance for individual tools was similarly strong: for the CT slice retrieval tool, the system achieved accuracy of 97.7% (96.5–98.7%) and F1 score of 0.906 (0.855–0.947); for the anatomical segmentation tool, the system achieved accuracy of 95.4% (93.8–96.8%) and F1 score of 0.878 (0.834–0.917); for no tool call, the system achieved accuracy of 93.4% (648 of 694; 91.5–95.1%) and F1 score of 0.952 (0.937–0.965).

Variations in planner prompting strategies had a greater impact on tool-calling accuracy compared to variations in executor prompting strategies, highlighting the planner’s critical role in guiding accurate tool invocation. Notably, the few-shot planner paired with a zero-shot executor achieved a comparable overall tool-calling accuracy to the few-shot planner-executor of 92.1% (639 of 694; 90.1–94.1%). In contrast, the zero-shot planner variations performed slightly worse, with the zero-shot planner and few-shot executor yielding an overall tool-calling accuracy of 89.0% (618 of 694; 86.6–91.4%), and zero-shot prompting for both agents resulting in the lowest overall tool-calling accuracy of 88.8% (616 of 694; 86.3–91.1%).

### LLM-as-a-judge for Self-Regulation

The agentic system’s self-regulation ability to appropriately defer tool-independent questions beyond its capabilities was evaluated using an LLM-as-a-judge. Results are summarized in Table 4. As with tool-calling accuracy, all variations of the planner-executor architecture achieved significantly higher overall self-regulation accuracy compared to the single-agent baseline (all P < .001), which achieved an accuracy of 64.7% (323 of 499; 60.5–68.9%). These findings emphasize the critical role of multi-agent architecture in enabling effective self-regulation.

**Table 4:** Self-regulation accuracy with varying prompting strategies in the planner and executor agents.

| Planner Prompt | Executor Prompt | Classification | Precision (%)<br>(95% CI) | Recall (%)<br>(95% CI) | F1 Score<br>(95% CI) | Overall Accuracy (%)<br>(95% CI) | Adjusted P-value<sup>a</sup> |
| --- | --- | --- | --- | --- | --- | --- | --- |
| Few-shot | Few-shot | Answerable | 88.2 (270/306)<br>(85.4–91.7) | 99.6 (270/271)<br>(98.8–100) | 0.936<br>(0.914–0.955) | 92.6 (462/499)<br>(90.2–94.8) | <0.001 |
|  |  | Defer-to-doctor | 99.5 (192/193)<br>(98.3–100) | 84.2 (192/228)<br>(79.4–88.6) | 0.912<br>(0.882–0.938) |  |  |
| Zero-shot | Few-shot | Answerable | 82.1 (271/330)<br>(77.7–86.2) | 100 (271/271)<br>(100–100) | 0.902<br>(0.875–0.926) | 88.2 (440/499)<br>(85.2–91.0) | <0.001 |
|  |  | Defer-to-doctor | 100 (169/169)<br>(100–100) | 74.1 (169/228)<br>(68.4–79.7) | 0.851<br>(0.813–0.887) |  |  |
| Few-shot | Zero-shot | Answerable | 87.5 (267/305)<br>(83.7–91.2) | 98.5 (267/271)<br>(97.0–99.7) | 0.927<br>(0.904–0.948) | 91.6 (457/499)<br>(89.0–94.0) | <0.001 |
|  |  | Defer-to-doctor | 97.9 (190/194)<br>(95.8–99.5) | 83.3 (190/228)<br>(78.2–88.0) | 0.900<br>(0.868–0.929) |  |  |
| Zero-shot | Zero-shot | Answerable | 80.2 (268/334)<br>(75.9–84.4) | 98.9 (268/271)<br>(97.5–100) | 0.886<br>(0.858–0.911) | 86.2 (430/499)<br>(83.0–89.0) | <0.001 |
|  |  | Defer-to-doctor | 98.2 (162/165)<br>(95.8–100) | 71.1 (162/228)<br>(65.0–76.9) | 0.824<br>(0.781–0.864) |  |  |
| Single-agent baseline |  | Answerable | 61.0 (263/431)<br>(56.4–65.5) | 97.0 (263/271)<br>(94.8–98.9) | 0.749<br>(0.713–0.783) | 64.7 (323/499)<br>(60.5–68.9) | N/A |
|  |  | Defer-to-doctor | 88.2 (60/68)<br>(79.7–95.4) | 26.3 (60/228)<br>(20.7–32.2) | 0.405<br>(0.333–0.476) |  |  |

a: McNemar’s test with Bonferroni correction for multiple comparisons was used to compare accuracy of multi-agent variations against the single-agent baseline, with adjusted significance level $P \leq 0.0125$ ( $\alpha=.05/4$ ).

Self-regulation accuracy patterns mirrored those observed with tool-calling accuracy. The planner-executor system using few-shot prompting for both agents achieved an overall self-regulation accuracy of 92.6% (462 of 499; 90.2–94.8%). For answerable questions, the system achieved precision of 88.2% (270 of 306; 85.4–91.7%), recall of 99.6% (270 of 271; 98.8–100%), and F1 score of 0.936 (0.914–0.955). For defer-to-doctor questions, the system achieved precision of 99.5% (192 of 228; 98.3–100%), recall of 84.2% (192 of 228; 79.4–88.6%), and F1 score of 0.9122 (0.882–0.938).

Variations in planner prompting strategies had a greater impact on overall self-regulation accuracy compared to executor prompting strategies, further emphasizing the planner’s critical role in self-regulation. The few-shot planner paired with a zero-shot executor achieved an overall self-regulation accuracy of 91.6% (457 of 499; 89.0–94.0%). In contrast, the zero-shot planner variations performed slightly worse, with the zero-shot planner and few-shot executor achieving an overall self-regulation accuracy of 88.2% (440 of 499; 85.2–91.0%), and zero-shot prompting for both agents resulting in the lowest overall self-regulation accuracy of 86.2% (430 of 499; 83.0–89.0%). Additional results for both tool-calling and self-regulation for variations in model temperature are provided in Supplementary Fig. 7.

### Reader Performance Study

The reader performance study evaluated the clinical quality of responses generated by the few-shot planner-executor system, selected based on its superior performance in both tool-calling and self-regulation accuracy. For answerable questions, the system demonstrated consistently strong performance across evaluation criteria, with 78.9% (180 of 228) of responses receiving perfect 5.0 scores for clinical accuracy, 72.4% (165 of 228) for comprehensiveness, 58.3% (133 of 228) for conciseness, and 73.2% (167 of 228) for patient understandability across all reviewers. For defer-to-doctor questions, the system maintained high performance, with 77.3% (133 of 172) of responses achieving perfect 5.0 scores for clinical accuracy (P = 0.715 vs answerable), 70.3% (121 of 172) for relevance, 47.1% (81 of 172) for conciseness (P = 0.027 vs answerable), and 70.3% (121 of 172) for patient understandability (P = 0.574 vs answerable). Because the system was instructed to provide relevant clinical context prior to deferring to a provider, P = 0.027 vs. answerable in the percentage of perfect conciseness scores for defer-to-doctor questions is reflective of longer responses in this category despite accurate and relevant information retrieval. Fig. 3 illustrates the full distribution of reviewer ratings for both question types.

**Figure 3.**
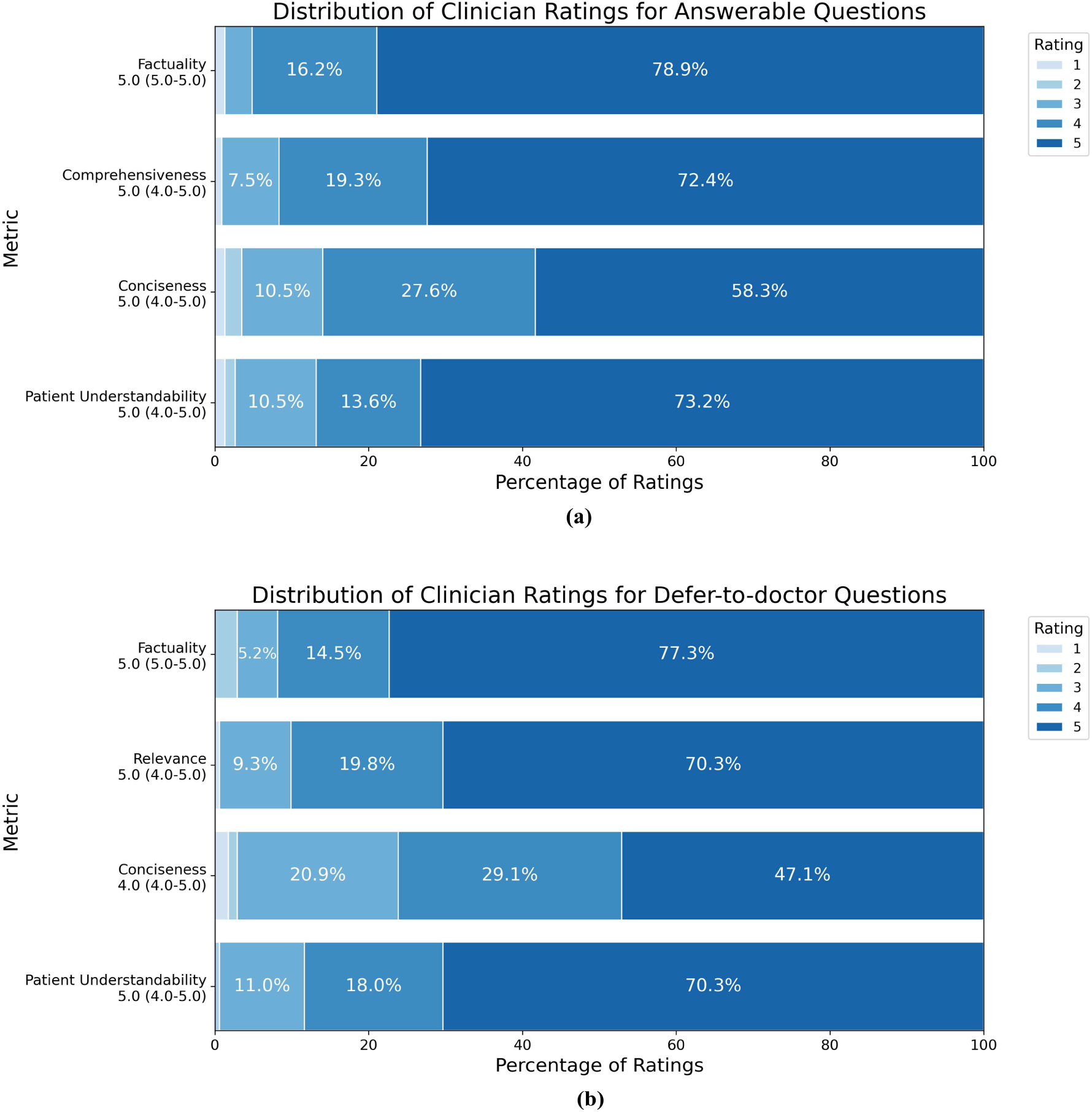
Distribution of clinician ratings across four readers for a subset of 100 agentic system responses evaluated in the reader performance study. (a) Answerable questions. (b) Defer-to-doctor questions. Responses were rated using a 1-5 Likert scale on factuality, comprehensiveness (answerable) or relevance (defer-to-doctor), conciseness, and patient understandability. Median ratings with interquartile range (IQR) are shown for each metric.

For answerable questions, readers showed high agreement with Gwet’s AC2 of 0.92 (SE=0.03) for factuality, 0.89 (0.08) for comprehensiveness, 0.81 (0.04) for conciseness, and 0.86 (0.03) for patient understandability. For defer-to-doctor questions, agreement was slightly lower with coefficients of 0.80 (SE=0.04) for factuality, 0.88 (0.15) for relevance, 0.73 (0.14) for conciseness and 0.78 (0.13) for patient understandability.

Sample system responses to all four question types are provided in Fig. 4.

**Figure 4.**
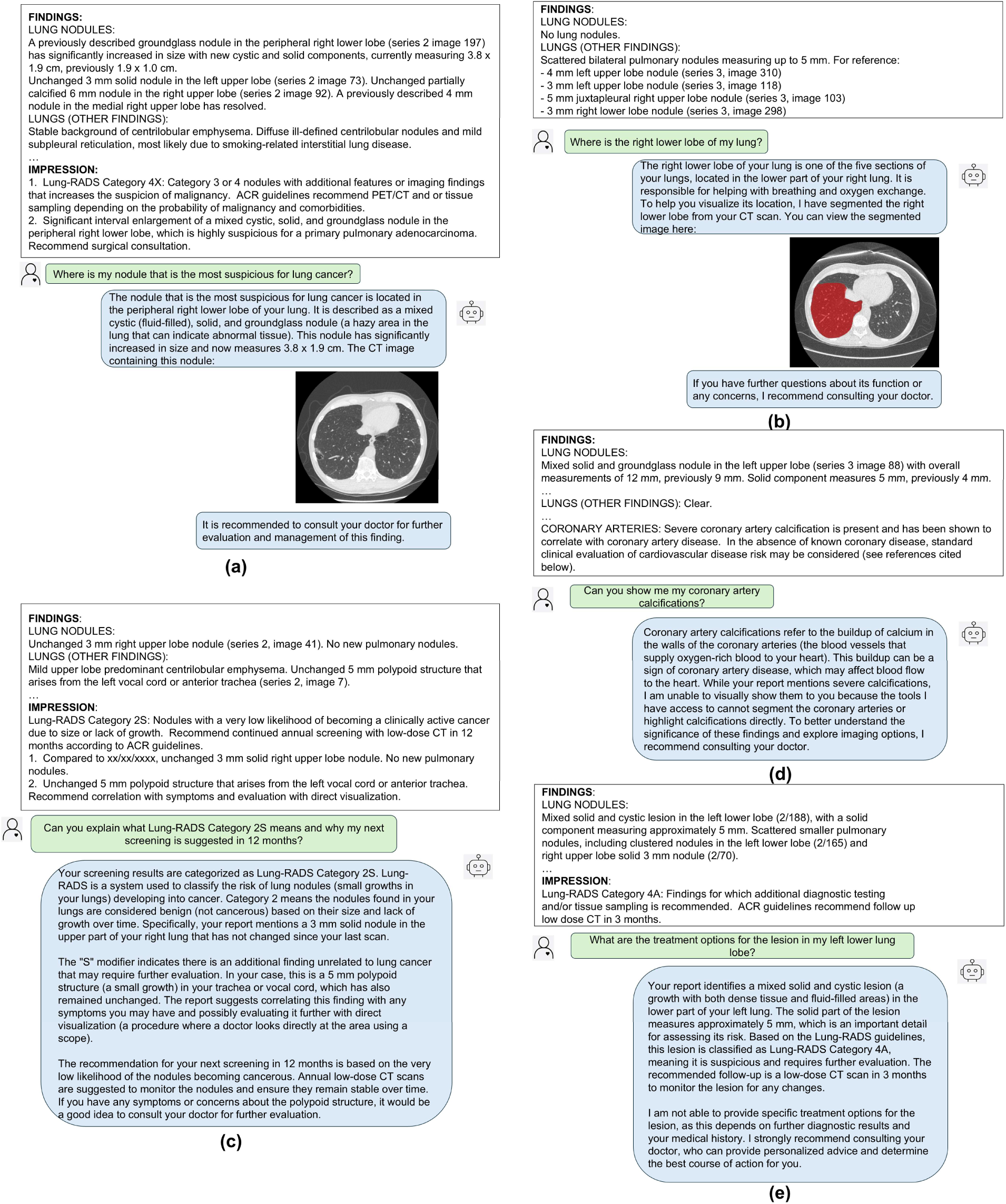
Sample agentic system responses. (a) Answerable image-based query requiring CT slice retrieval tool. (b) Answerable image-based query requiring segmentation tool. (c) Answerable text-based query asking for interpretation of Lung-RADS category. (d) Defer-to-doctor image-based query. (e) Defer-to-doctor text-based query asking about patient-specific treatment.

### Error Analysis

Errors made by the agentic system for tool-calling, self-regulation, and lowest-scoring responses in the reader performance study tended to be sporadic and are provided in Fig. 5. For tool-calling, common errors involved incorrectly calling the CT slice retrieval tool using an inaccurate series or image number Fig. 5(a) or segmenting unrelated anatomical structures Fig. 5(b). Notable errors in self-regulation included making absolute statements of patient-specific treatments Fig. 5(c). Additional notable errors included minor misinterpretation of Lung-RADS guidelines or related medical terms. Quantification of tool-calling errors, characterized by correct tool invocation but incorrect argument parameters, and low-scoring reader study responses are provided in Supplementary Fig. 8 and 9 respectively.

**Figure 5.**
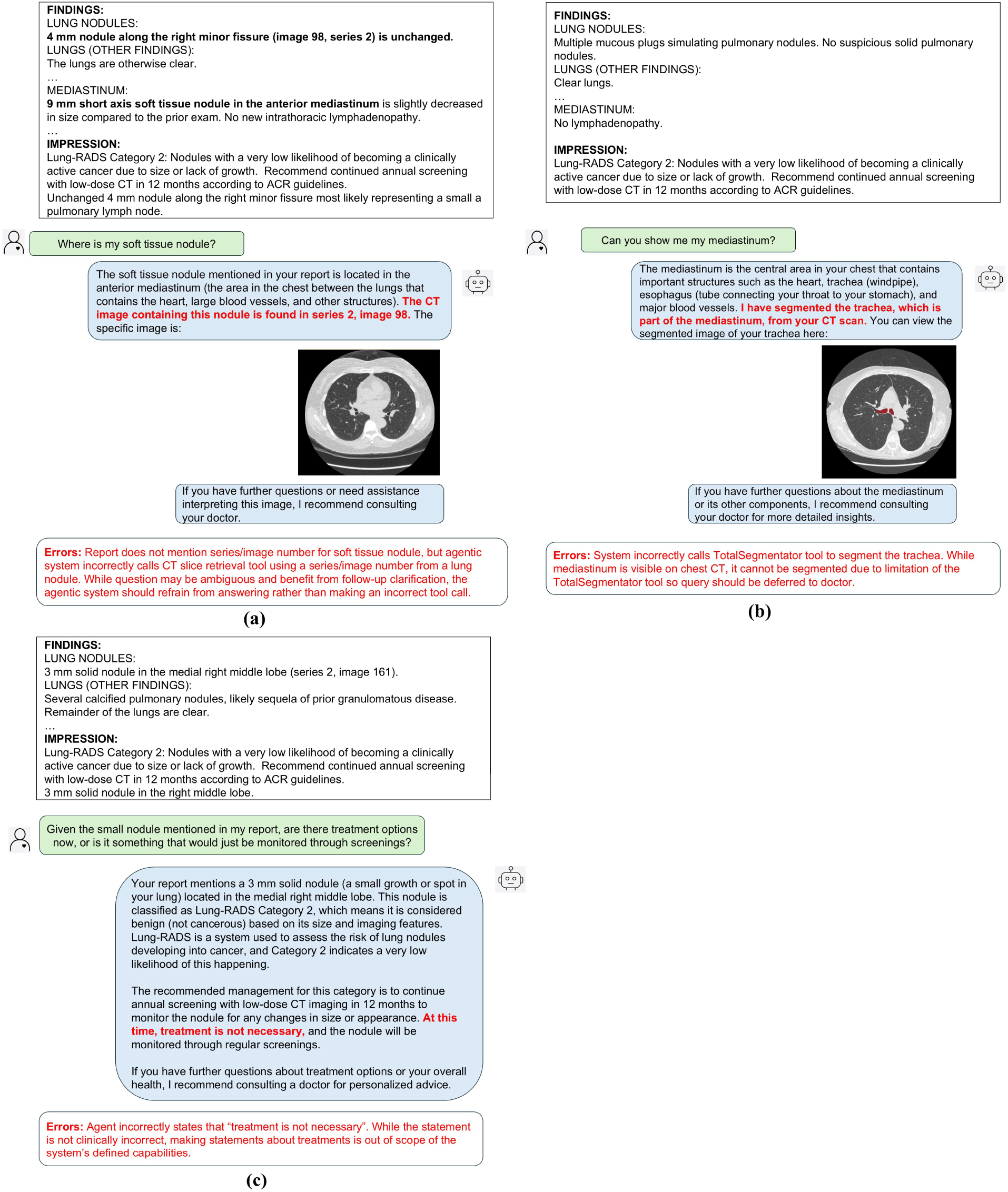
Sample incorrect agentic system responses. (a) Incorrect tool call to CT slice finder tool. (b) Incorrect tool call to segmentation tool. (c) System going out of scope of defined capabilities in a defer-to-doctor question.

## DISCUSSION

Effective communication of lung cancer screening (LCS) results is crucial to improving patient engagement to follow-up and screening, emphasizing the need for scalable solutions to enhance understanding and support timely intervention. In this study, we developed and evaluated a multimodal agentic AI system capable of answering simulated patient-specific questions about LCS CT results. The multi-agent system demonstrated high tool-calling and self-regulation accuracy, significantly outperforming the single-agent baseline. Reader study results further confirmed strong clinical accuracy and patient understandability across both answerable and defer-to-doctor questions.

An agentic system with clinical accuracy and self-regulation has significant potential for enhancing patient communication in LCS. Studies [22], [23] emphasize the importance of patient engagement and understanding in LCS to improve compliance and continued screening, one of the greatest barriers in LCS currently as evident in the low screening and follow-up adherence [1], [2]. Heightened patient interaction with medical records and interest in medical imaging reports following the 21st Century Cures Act [4] further highlights the need for scalable strategies that support patient comprehension. Our agentic system offers an advanced patient communication tool that in clinical practice could be integrated as a standalone tool with appropriate regulatory safeguards or alongside a radiologist or LCS team clinician to assist in addressing patient questions related to their screening results. While prior work has demonstrated the potential of native LLMs for tasks like patient-friendly report generation [5], [6], concept-based explanations [24], and answering general questions about LCS [25], these systems lack the ability to integrate specialized imaging data. Additionally, existing agentic systems in the medical domain have largely been focused on clinical decision-making or diagnostic tasks [11], [12].

Furthermore, our system’s ability to defer certain questions to physicians with high accuracy demonstrates effective self-regulation, which is essential in the medical domain as erroneous or unintended responses by AI could lead to confusion or heightened anxiety. This capability is particularly important in patient communication, especially in LCS, where reliable information is crucial to promote patient engagement and adherence to follow-up. Prior studies have explored evaluating regulatory capabilities of LLM-based systems both in the general language setting [26] and medical domain [27], with a focus on tasks such as clinical decision-making or diagnostic reasoning [28], [29]. Our work extends this to the patient communication setting. Additionally, prior work has demonstrated the effectiveness of multi-agent systems in mirroring clinical workflows within diagnostic and medical decision-making settings [12], [30], [31]. Building on this, we employ a planner-executor style for patient communication in LCS, enabling the system to effectively structure and execute responses that align with clinical reasoning.

This study had several limitations. First, while the set of simulated patient questions generated with LLM assistance were carefully designed with input from multiple clinicians to cover a diverse range of tasks and clinically relevant queries, they were not directly sourced from patients. Future work could expand this by collecting a broader range of questions directly from patients to better reflect the variety of queries possible in the real-world clinical setting. Additionally, the evaluation was limited in addressing ambiguous patient questions that might require further clarification due to the straightforward nature of the simulated queries. Future work could address this by incorporating follow-up capabilities to enable the system to engage in multi-turn conversations with patients. Furthermore, while the agentic system demonstrated strong performance in utilizing its current imaging tools as appropriate to answer patient queries, its capabilities could be expanded with additional tools such as nodule detection algorithms or advanced imaging techniques to improve personalized image interpretation. Lastly, while the Lung-RADS guidelines and imaging tools are specific to the LCS setting, future work could explore extending the planner-executor framework to other medical imaging domains and clinical communication workflows.

## CONCLUSION

We developed a multimodal agentic AI system capable of answering patient questions about lung cancer screening CT results, specifically designed with self-regulation capabilities, and evaluated the system across a diverse set of simulated patient queries regarding LCS. By demonstrating robust tool-calling accuracy, effective self-regulation, and strong performance in clinical accuracy and patient understandability, the agentic system highlights the potential for such systems to enhance patient communication in clinical workflows.

## Data Availability

The data that support the findings of this study are not publicly available due to institutional and patient privacy restrictions. The data may be made available upon reasonable request and with permission from the University of California, San Francisco (UCSF), subject to applicable institutional approvals and data-use requirements.

## Abbreviations

LCS: Lung Cancer Screening
LLM: Large Language Model
AI: Artificial Intelligence

## ACKNOWLEDGEMENTS

Ali Nowroozi, MD was an NIH T32 postdoctoral fellow for most of this project, under the award number 5T32HL007185-47. A.S. Rustagi states: the opinions expressed herein are those of the authors and not their employers, the U.S. Department of Veterans Affairs, the U.S. government, or the study sponsors. Figures 1, 2, 4, 5 contain: “doctors” icon by SANB, "tool" by rendicon, “guidelines” icon by Muhammad Atiq, “ai assistant” icon by Anamika singh, “execution” by Nithinan Tatah, “Patient” by Mila Yunita from thenounproject.com CC BY 3.0.

## APPENDIX

## A. Appendix 1.1

Statistical power analysis determined that at least 75 questions were necessary per task to calculate per-tool call accuracy with a 95% confidence interval (±5%). One component of the evaluation was assessing tool-calling accuracy of the CT slice retrieval tool, which required mention of a finding localized with exact series and image number referenced in the report. This was present in approximately 65% of the initial set of reports prior to sampling. Based on this estimation, the sample size was iteratively adjusted to ensure at least 75 reports containing a finding with series and image number reference were included. This process resulted in the selection of 116 reports, of which 79 contained the required localization reference, exceeding the minimum threshold. For other components of the evaluation, the same 75-report requirement was applied to ensure sufficient data for analysis. However, these components did not require iterative adjustments, as the selected subset of 116 reports already included more than 75 reports meeting their criteria.

## B. Appendix 1.2

Stratified sampling based on retrospective analysis of clinical demographics at our institution was performed to ensure accuracy in Lung-RADS category distribution for the LCS reports used to generate the simulated patient questions. Lung-RADS categories were distributed as follows: 33.7% of cases in Lung-RADS-1, 56.1% in Lung-RADS-2, 4.8% in Lung-RADS-3, 2.3% in Lung-RADS-4A, 1.2% in Lung-RADS-4B, and 0.7% in Lung-RADS-4X. Report samples were proportionally allocated to meet these quotas, with random selection performed within each category.

## REFERENCES

[1] R. Y. Kim et al., “Adherence to Annual Lung Cancer Screening and Rates of Cancer Diagnosis,” JAMA Netw Open, vol. 8, no. 3, p. e250942, Mar. 2025, doi: 10.1001/jamanetworkopen.2025.0942.

[2] A. T. Gudina et al., “Lung Cancer Screening Uptake under the Revised United States Preventive Services Task Force Guideline: Assessing Disparities,” Cancer Epidemiol Biomarkers Prev, vol. 34, no. 1, pp. 35–41, Jan. 2025, doi: 10.1158/1055-9965.EPI-24-0725.

[3] D. Alomar, M. Almashmoum, I. Eleftheriou, P. Whelan, and J. Ainsworth, “The Impact of Patient Access to Electronic Health Records on Health Care Engagement: Systematic Review,” J Med Internet Res, vol. 26, p. e56473, Nov. 2024, doi: 10.2196/56473.

[4] J. R. Pollock, S. A. B. Petty, J. J. Schmitz, J. Varner, A. M. Metcalfe, and N. Tan, “Patient Access of Their Radiology Reports Before and After Implementation of 21st Century Cures Act Information-Blocking Provisions at a Large Multicampus Health System,” AJR Am J Roentgenol, vol. 222, no. 6, p. e2330343, Jun. 2024, doi: 10.2214/AJR.23.30343.

[5] S. Bannur, et al., “MAIRA-2: Grounded Radiology Report Generation,” Sep. 20, 2024, arXiv: arXiv:2406.04449. doi: 10.48550/arXiv.2406.04449.

[6] J. Park, K. Oh, K. Han, and Y. H. Lee, “Patient-centered radiology reports with generative artificial intelligence: adding value to radiology reporting,” Sci Rep, vol. 14, no. 1, p. 13218, Jun. 2024, doi: 10.1038/s41598-024-63824-z.

[7] T. Meeradevi, S. Sasikala, L. Murali, N. Manikandan, and K. Ramaswamy, “Lung cancer detection with machine learning classifiers with multi-attribute decision-making system and deep learning model,” Sci Rep, vol. 15, no. 1, p. 8565, Mar. 2025, doi: 10.1038/s41598-025-88188-w.

[8] D. Ardila et al., “End-to-end lung cancer screening with three-dimensional deep learning on low-dose chest computed tomography,” Nat Med, vol. 25, no. 6, pp. 954–961, Jun. 2019, doi: 10.1038/s41591-019-0447-x.

[9] J. Christensen, et al., “ACR Lung-RADS v2022: Assessment Categories and Management Recommendations,” Journal of the American College of Radiology, vol. 21, no. 3, pp. 473–488, Mar. 2024, doi: 10.1016/j.jacr.2023.09.009.

[10] OpenAI, et al., “GPT-4o System Card,” Oct. 25, 2024, arXiv: arXiv:2410.21276. doi: 10.48550/arXiv.2410.21276.

[11] A. Fallahpour, J. Ma, A. Munim, H. Lyu, and B. Wang, “MedRAX: Medical Reasoning Agent for Chest X-ray,” Feb. 04, 2025, arXiv: arXiv:2502.02673. doi: 10.48550/arXiv.2502.02673.

[12] Z. Wang, et al., “MedAgent-Pro: Towards Evidence-based Multi-modal Medical Diagnosis via Reasoning Agentic Workflow,” Jul. 02, 2025, arXiv: arXiv:2503.18968. doi: 10.48550/arXiv.2503.18968.

[13] J. Wasserthal et al., “TotalSegmentator: Robust Segmentation of 104 Anatomic Structures in CT Images,” Radiology: Artificial Intelligence, vol. 5, no. 5, p. e230024, Sep. 2023, doi: 10.1148/ryai.230024.

[14] T. Kojima, S. S. Gu, M. Reid, Y. Matsuo, and Y. Iwasawa, “Large Language Models are Zero-Shot Reasoners,” Jan. 29, 2023, arXiv: arXiv:2205.11916. doi: 10.48550/arXiv.2205.11916.

[15] T. B. Brown, et al., “Language Models are Few-Shot Learners,” Jul. 22, 2020, arXiv: arXiv:2005.14165. doi: 10.48550/arXiv.2005.14165.

[16] S. Rajagopal, J. H. Sohn, H. Subramonyam, and S. Mohan, “Can Generative AI Support Patients’ & Caregivers’ Informational Needs? Towards Task-Centric Evaluation Of AI Systems,” Feb. 28, 2025, arXiv: arXiv:2402.00234. doi: 10.48550/arXiv.2402.00234.

[17] H. Li, et al., “LLMs-as-Judges: A Comprehensive Survey on LLM-based Evaluation Methods,” Dec. 11, 2024, arXiv: arXiv:2412.05579. doi: 10.48550/arXiv.2412.05579.

[18] L. Zheng, et al., “Judging LLM-as-a-Judge with MT-Bench and Chatbot Arena,” Dec. 24, 2023, arXiv: arXiv:2306.05685.doi: 10.48550/arXiv.2306.05685.

[19] Q. McNemar, “Note on the Sampling Error of the Difference Between Correlated Proportions or Percentages,” Psychometrika, vol. 12, no. 2, pp. 153–157, Jun. 1947, doi: 10.1007/BF02295996.

[20] K. L. Gwet, Handbook of inter-rater reliability: the definitive guide to measuring the extent of agreement among raters, Fourth edition. Gaithersburg, Md: Advances Analytics, LLC, 2014.

[21] R. A. Fisher, “On the Interpretation of χ 2 from Contingency Tables, and the Calculation of P,” Journal of the Royal Statistical Society, vol. 85, no. 1, p. 87, Jan. 1922, doi: 10.2307/2340521.

[22] J. Jung et al., “Patient-Reported Barriers and Preferred Interventions to Improve Lung Cancer Screening Uptake,” Journal of the American College of Radiology, vol. 22, no. 3, pp. 269–279, Mar. 2025, doi: 10.1016/j.jacr.2024.10.010.

[23] Y. Yang, W. Zhao, D. N. S. Chan, S. Zhou, K. C. Choi, and W. K. W. So, “Interventions targeted to improve lung cancer screening uptake among high-risk individuals: A systematic review and meta-analysis,” Asia Pac J Oncol Nurs, vol. 12, p. 100746, Jun. 2025, doi: 10.1016/j.apjon.2025.100746.

[24] S. E. Herwald, P. Shah, A. Johnston, C. Olsen, J.-B. Delbrouck, and C. P. Langlotz, “RadGPT: A System Based on a Large Language Model That Generates Sets of Patient-Centered Materials to Explain Radiology Report Information,” Journal of the American College of Radiology, vol. 22, no. 9, pp. 1050–1059, Sep. 2025, doi: 10.1016/j.jacr.2025.06.013.

[25] H. L. Haver, C. T. Lin, A. Sirajuddin, P. H. Yi, and J. Jeudy, “Use of ChatGPT, GPT-4, and Bard to Improve Readability of ChatGPT’s Answers to Common Questions About Lung Cancer and Lung Cancer Screening,” American Journal of Roentgenology, vol. 221, no. 5, pp. 701–704, Nov. 2023, doi: 10.2214/AJR.23.29622.

[26] Y. Deng, Y. Zhao, M. Li, S.-K. Ng, and T.-S. Chua, “Don’t Just Say ‘I don’t know’! Self-aligning Large Language Models for Responding to Unknown Questions with Explanations,” Oct. 02, 2024, arXiv: arXiv:2402.15062. doi: 10.48550/arXiv.2402.15062.

[27] C.-K. Wu, Z. R. Tam, C.-Y. Lin, Y.-N. Chen, and H. Lee, “Answer, Refuse, or Guess? Investigating Risk-Aware Decision Making in Language Models,” Mar. 03, 2025, arXiv: arXiv:2503.01332. doi: 10.48550/arXiv.2503.01332.

[28] M. Griot, C. Hemptinne, J. Vanderdonckt, and D. Yuksel, “Large Language Models lack essential metacognition for reliable medical reasoning,” Nat Commun, vol. 16, no. 1, p. 642, Jan. 2025, doi: 10.1038/s41467-024-55628-6.

[29] A. Pal, L. K. Umapathi, and M. Sankarasubbu, “Med-HALT: Medical Domain Hallucination Test for Large Language Models,” Oct. 14, 2023, arXiv: arXiv:2307.15343. doi: 10.48550/arXiv.2307.15343.

[30] X. Tang, et al., “MedAgents: Large Language Models as Collaborators for Zero-shot Medical Reasoning,” Jun. 04, 2024, arXiv: arXiv:2311.10537. doi: 10.48550/arXiv.2311.10537.

[31] Y. Kim, et al., “MDAgents: An Adaptive Collaboration of LLMs for Medical Decision-Making,” Oct. 30, 2024, arXiv: arXiv:2404.15155. doi: 10.48550/arXiv.2404.15155.

